# Examining gender and ethnic disparities in scientific authorship to promote a culture of equity, diversity and inclusion at a university school of public health

**DOI:** 10.1101/2025.02.01.25321228

**Authors:** Paula Christen, Julia Michalow, Tristan Naidoo, Hillary Topazian, Sabine van Elsland, Abeer M Arif, Marc Baguelin, Gemma Clunie, Daniela Fecht, Tini Garske, Sondus Hassounah, Jenny Husbands, Wendy Kwok, Sequoia I Leuba, Clare McCormack, Kate M Mitchell, Matteo Pianella, Michael Pickles, Shazia Ruybal-Pesántez, Nora Schmit, Chi Udeh-Momoh, Sarah Essilfie-Quaye, Anne Cori, Isobel M Blake, Lucy Okell

## Abstract

While fostering equity, diversity, and inclusion in the workplace is crucial for many reasons, in public health research, diverse perspectives are particularly vital to identify biases that homogenous teams might miss. Given that publication metrics often influence career progression, we investigated gender and ethnic disparities in publication rates within Imperial College London’s School of Public Health.

We analysed 18,322 peer-reviewed publications by 513 affiliated researchers between 2014 and 2023. We used mixed-effects multivariable regression models to assess the impact of gender, ethnicity, and job level on publication rates. Network analysis of co-authorships was performed to evaluate each author’s centrality in the department’s research network.

We found a persistent gender gap in publication rates across all job levels and ethnicities, with men publishing more than women (incidence rate ratio (IRR) 1.30 95% Confidence Interval (CI): 1.15 – 1.46). This disparity was present from early career levels and amplified in senior roles, where men were overrepresented (71.2% of men at Professor level). Internal collaboration rates were similar between genders.

Unadjusted analyses indicated higher publication rates for white researchers (median of 1 publication more per person per year), although there were limitations in the ethnicity classification algorithm.

The COVID-19 pandemic led to increased publication rates for both genders, but the gender gap persisted, with men publishing 1.27 (95% CI: 1.10 – 1.46) times more than women in 2020 - 2021. A complex interplay of factors may contribute to publication disparities, including differences in research contributions, systemic barriers, and potential biases in research allocation, mentorship, and promotion processes.

This study underscores the need to identify and address the root causes of these disparities to foster an inclusive research environment where diverse contributions are recognized and valued.

## Background

Reasons for fostering equity, diversity and inclusion (EDI) in the workplace are vast. In public health research, diverse perspectives enhance the ability to identify biases that homogenous teams would otherwise overlook. ^1,2^ Increasing diversity within research teams can enhance the quality of research, and help address the unique public health challenges faced by different population subgroups.^3^ Beyond this, we need to act on the systemic issues in education, academia and the scientific publishing community that have led to inequities and unequal opportunities.

Despite progress driven by EDI initiatives such as the Athena Swan Charter^4^ and Race Equality Charter in the UK^5^, evidenced by high entry rates for pupils from Chinese, Indian, and Black African backgrounds into higher education^6^, inequities persist across the educational and publishing landscape. Growing concerns about boys being left behind in school are mirrored in higher education^7^, where women now outnumber men in enrolment.^8^ Yet, women remain underrepresented in the upper echelons of academia, which are dominated by white men. ^9^ This phenomenon, often referred to as the “leaky pipeline” ^9^ or “scissors effect” ^10^, is well-documented across many disciplines and countries. ^11–15^

Despite the importance of understanding representation in academia, the position of minority ethnic groups in academia has received little attention.^16^ In the United States (US), women from minoritized groups are significantly underrepresented in science, technology, engineering, and mathematics (STEM) faculty compared to their share of the population, and this disparity worsens at higher academic levels.^17,18^ Faculty diversity is vital, as it can significantly reduce performance gaps between students of different racial and ethnic backgrounds.^17^ Moreover, a significant proportion of minority students^19^ as well as faculty members^20^ report experiencing discrimination or harassment, highlighting the urgent need to address systemic inequities in academia.

Gender bias has been identified in teaching ratings and salaries^21^, two critical factors for retaining staff in academia.^22,23^ However, contrary to omnipresent claims in the literature of sexism in the tenure-track academy ^24–28^, a review of the literature on gender bias in academia from 2000 to 2020 across multiple faculties in institutions across the US^21^ surprisingly concludes that, while there is lower representation of women across job categories in mathematically intensive fields, overall tenure-track women are on par with tenure-track men in grant funding and journal acceptance rates. Yet, these findings vary across disciplines and may differ globally.^29–32^ However, few studies have accounted for observed productivity, which can moderate the tenure track evaluation contexts.

Publication count, an incomplete quantification of productivity, is arguably a main currency of the academic profession, and a key metric considered in promotions.^33–35^ Yet, the reality of productivity is more broad, encompassing not only traditional tangible research output (such as publication count) but also time devoted to teaching, community engagement, and “academic housework” – activities often disproportionately undertaken by women^36^ yet rarely formally acknowledged in promotion processes. ^37,38^ Nygaard et al. demonstrated the disproportionate benefits men receive from using publication output and impact as primary productivity indicators.^39^ The need for a more comprehensive measure of productivity in academia has been recognized ^40^, but there are no standard guidelines on accounting for the full array of activities to allow for comparable promotion processes across institutions or even faculties. ^34^

Research exploring publication disparities in academia, especially those related to ethnicity, race, and the intersectionality of these factors with gender, remains limited. Existing studies have revealed concerning trends: Black and Hispanic medical school graduates in the US publish significantly less than their Asian and White counterparts^41^, while large-scale analyses have identified persistent gender gaps in publication count and first and last authorship^32,42–44^, though some evidence suggests these may be decreasing.^29,43^ However, few studies have comprehensively examined these biases across various job levels within faculty departments or isolated the specific impacts of ethnicity and race, particularly when interconnected with gender. Much of the existing work focuses on authorship within journals rather than the broader context of departmental productivity. In addition, most work that tackles the productivity causality puzzle focuses solely on the US ^21^, specific medical domains/sub-fields^45^, or compares multiple disciplines. ^39,46^

We have conducted research to contribute to specific takes on this issue by focusing on the field of public health and examining the intersection of gender and ethnicity in the context of publication productivity in the United Kingdom. We sought to understand whether publication rates contribute to the leaky pipeline, resulting in gender and ethnic disparity in our department, the School of Public Health (SPH), Imperial College London (ICL). Supported by a broader working group, our core writing team comprises* and represents views from PhD student to Reader level. Ultimately, this study contributes to a growing body of evidence advocating for tailored evaluation methods that acknowledge the diverse research practices and publication norms across different fields and regions, promoting a more equitable academic environment. In this commentary, we provide an overview of our findings, discuss potential explanations for the findings and propose avenues for fostering a more equitable and inclusive environment within similar departments.

## Methods

We conducted a quantitative analysis of peer-reviewed papers published by researchers within SPH (Figure 1). This department comprises researchers in Epidemiology and Biostatistics, Infectious Disease Epidemiology, Primary Care and Public Health, Imperial Clinical Trials Unit, Ageing Epidemiology Research Unit and the Environmental Research Group. We quantified the publication rate by gender and ethnicity over time, adjusting for job level. Publications were obtained from Symplectic Elements^47^ (the research information management system used by ICL) and gender was assigned using an algorithm based on historical name datasets and manual review. ^48^ We contrast “man” and “woman” in this piece due to lack of data on gender identity, but acknowledge the existence of non-binary, intersex, and other diverse gender identities. Job categories were scraped from ICL staff webpages. As information on promotion dates were not readily available, we relied on the assumption that the most recent SPH job title was an individual’s job category across all publication records. Ethnicity was assigned using probabilistic estimates based on full names and US Census data. ^49^ The number of individuals estimated to be from different ethnicities were low such that we categorized White individuals as “non-minoritized” and Asian, Black and Hispanic individuals as “minoritized” to ensure individual privacy. While this ethnicity categorization is likely to be inaccurate in many cases, studies have shown substantial unconscious bias can result from a name alone (e.g., in controlled recruitment studies based on CVs).^50^ Therefore we considered that ‘perceived ethnicity’ may still be a useful indicator of potential for bias for example from journal editors or reviewers when a researcher submits their paper – even if it does not always reflect a person’s lived experience at work.

**Figure 1.**
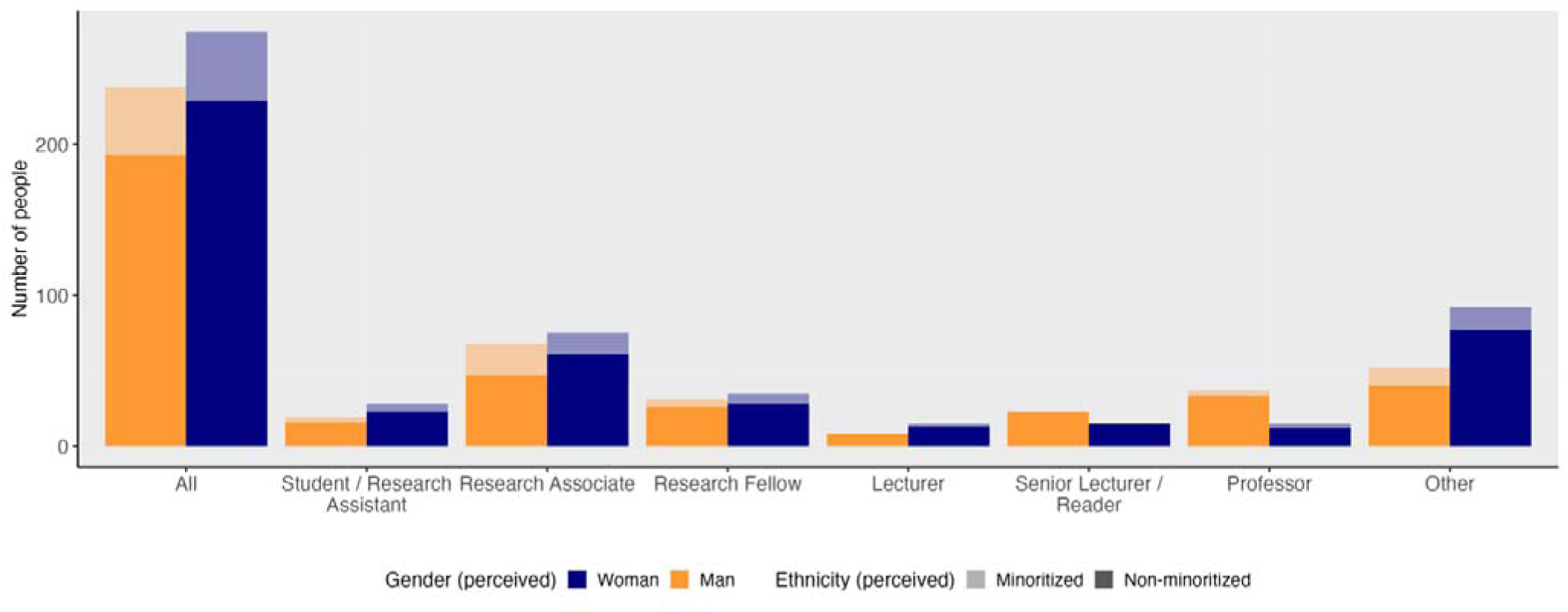
Proportion of men and women researchers by job category and perceived ethnicity group at the School of Public Health, Imperial College London, 2014-2023. See Data Collection (job category) section for more information on “other”.

Negative binomial mixed-effects regression models were used to assess the effects of gender (man, woman), ethnicity (minoritised, non-minoritised), job category (Student / Research Assistant, Research Associate, Research Fellow, Lecturer, Senior Lecturer / Reader, Professor, Other), and time period (2014/15, …, 2022/23) on annual number of publications per person, using random intercepts for each person. A network analysis was performed to quantify author centrality, measured in number of internal co-authors per year, within the SPH’s publication network. A more detailed account of the methods is presented in the Supplement.

## Results

To give a better sense of SPH, among the 275 women and 238 men researchers affiliated to the SPH between 2014 and 2023, there are more women than men from Student / Research Assistant (59.6% to 40.4%) to Lecturer level (65.2% to 34.8%), but men outnumber women in all more senior job categories (Senior Lecturers / Readers (39.5% to 60.5%), Professors (28.8% to 71.2%)) (Supplement, Figure 1). We observe a persistent gender gap in publication rates when analysing 18,322 papers by researchers affiliated to SPH between 2014 and 2023. Overall men publish a median of 2 (interquartile range (IQR): 1 – 4) papers per person per year, while women publish a median of 1 (IQR: 1 – 3) papers per person per year. This difference is more pronounced in senior job categories (Lecturers and Professors) (Supplement, Table 1).

Assuming that researchers are active every year between their first and last publication, we find that, on average, females in junior positions (Research Fellow and below) and senior positions (Lecturer and above) have career durations that are 8.1% and 9% shorter, respectively, than their male counterparts. We exclude the “Other” job category because it includes both junior and senior positions. Using unadjusted incidence rate ratios (IRR), we observe that, over the study period, men in junior positions achieved the same number of publications as women 4.27 years sooner, and 4.74 years sooner for men compared to women in senior positions.

We further investigated the difference in publication rates by adjusting for job category, ethnicity, and time period. We observed that men publish 1.30 (95% Confidence Interval (CI): 1.15 – 1.46) times more papers per person annually than women overall, and 1.31 (95% CI: 1.12 – 1.53) times more first or last author papers (Supplement, Table 1). The higher number of men at more senior job levels did not explain the difference in publication rates, with male Students / Research Assistants, Research Associates and Fellows publishing between 1.43 (95% CI: 0.89 – 2.29) and 1.34 (95% CI: 0.99 – 1.83) respectively more papers than their women counterparts. Men Senior Lecturers and Readers publish 1.68 (95% CI: 1.16 – 2.41) times more papers per person annually than their women counterparts. This trend persists among minoritised and non-minoritised researchers, with men consistently publishing more in both groups (Supplement, Table 2). The number of internal collaborations does not explain differences in publication rates by gender. Men, compared to women, had a similar average number of co-authors within the SPH (IRR = 1.01, 95% CI: 0.82 – 1.25).

Between 2014 and 2023, annual publication rates per researcher increased, especially during the COVID-19 pandemic, i.e., from 2020 onward. In 2020 and 2021, men and women published 1.17 (95% CI: 1.08 – 1.28) times more papers per person overall than in 2014 and 2015. For women, this trend picked up from 2018 onwards, while men’s annual publication rates increased from 2014 to 2019 and then decreased. Nonetheless, men continuously published significantly more than women from 1.29 (95% CI: 1.08 – 1.54) times more in 2014 and 2015 to 1.58 (95% CI: 1.35 – 1.86) times more in 2018 and 2019 (Supplement, Table 3). This gap reduced with men publishing 1.27 (95% CI: 1.10 – 1.46) and 1.07 (95% CI: 0.92 – 1.25) times more than women in 2020 and 2021, and 2022 and 2023 respectively.

Most researchers (82.3%) in SPH were classified as white by the ethnicity algorithm, although human resource data suggest the algorithm underestimates the proportion of minoritized staff based on name. The proportion in the ethnically minoritized group is consistent across all job levels and genders (Supplement, Figure 1). In the unadjusted analysis, researchers from minoritized groups published a median of 1 (IQR: 1 – 3) paper per person per year whereas researchers from non-minoritized groups published a median of 2 (IQR: 1 – 3) papers per person per year (Supplement, Figure 3). In the adjusted analysis we found no statistically significant difference in overall publication rates between researchers from minoritized and non-minoritized groups. However, among women, there was a marginally lower publication rate among ethnic minorities (minoritized women published 0.79 as many papers as non-minoritized women; 95% CI 0.63 – 1.00).

Men researchers from minoritized groups published significantly less as first or last authors compared to their men counterparts in non-minoritized groups (IRR= 0.83, 95% CI: 0.70 – 0.98), with a similar though non-significant trend among women researchers from minoritized groups (IRR= 0.79, 95% CI: 0.57 – 1.09). Authors from non-minoritized groups had a higher number of internal co-authors than researchers from minoritized groups in SPH (IRR= 1.81, 95% CI: 1.35 – 2.42).

## Discussion

We found a persistent gender gap in publication rates, with men publishing significantly more than women even after adjusting for job category, ethnicity, and time. These findings suggest that the higher publication rates and higher rates of first/last authorship among men researchers likely contribute to their overrepresentation in senior positions within SPH, since publication is a criterion for promotion.

Differences in publication output are often attributed to gender-specific parenthood roles, which disproportionately impact women’s career duration through factors such as maternity and part-time work.^51,52^ We hypothesized that these differences would manifest more prominently in senior positions, where career interruptions due to parenthood are more likely.^51^ After adjusting for differences in career length and job seniority, we found a 4.27-year gap in publishing between males and females in junior positions, with only a six month increase for senior positions. Assuming equal productivity during periods of full-time work, the observed gap appears too large to be explained by these factors alone.^53^

Our findings resonate with observations in Norway^39,54^ and Canada^55^, where multidisciplinary studies revealed men publish 20% to 40% more than women, highlighting that publication bias is a pervasive, potentially global issue demanding more attention. The gravity and breadth of this problem is underscored by the Declaration on Research Assessment (to which ICL has committed to since 2017) and the Leiden Manifesto, guidelines and recommendations crystallized at conferences held in 2012 and 2014^56^ respectively, which explicitly advise against using quantitative evidence on publication counts and impact only for individual researcher evaluations.

However, productivity in terms of publication count, does not necessarily reflect the time spent conducting the research or research engagement^57^, nor does it necessarily correlate with research complexity or research quality. Alternative metrics considered for promotions in research can be determined through a bibliometric approach (e.g., citation counts, citation rates, h-index); while these more accurately measure scholarly impact, they are correlated with publication count, and, thus, also subject to the gender and potential ethnicity biases we identified in our analysis. The inevitable consequence is the emphasis on quantity over quality, and the reinforcement of existing prestige hierarchies, potentially further disadvantaging women and minoritized researchers, known as the “Matthew effect”. ^39,58–61^

Our study revealed an increase in publication rates during the COVID-19 pandemic, particularly among women. This is contrary to other fields and faculties in other countries where women, especially Black women and mothers^62^, were negatively affected by the pandemic in terms of publication productivity. ^63,64^ This finding may be explained by the fact that many researchers in our departments had a “key worker” status during the pandemic, and thus had less caretaking responsibilities than researchers in other disciplines. But this finding also highlights the importance of conducting studies in different fields where specific contexts affect gender bias differently.

Although we have identified biases in publications within SPH, the complete set of reasons causing this bias is not immediately apparent. There is reasonable evidence that gender bias may exist in scientific publishing.^65^ While some studies show the chance of being published when submitting a paper to a journal is equal for all genders^46^, Kern-Goldberger’s^65^ and others’ studies demonstrate a difference in the acceptance rate based on perceived author. Holman et al propose that this could partially be explained by journals inviting men to submit papers at double the rate of women.^66^ However, the persistent gender gap suggests the issue extends beyond biases in the peer-review process. Long-standing claims attribute productivity gaps to caretaking responsibilities (maternity leave and caring for children and aging parents)^67^, with women having shorter career length^68^ and higher drop-out rates.^68,69^ But the literature suggests a more complex picture. Disparities in lifetime academic productivity can be traced to factors like unequal research funding^70^ and under- representation on editorial boards.^71^ Moreover, women disproportionally engage in teaching, community engagement, and “academic housework”^36^, possibly because they do not say “no”.^72^ Thus, evaluations ought to be more comprehensive to account for the various facets of activities researchers engage in, such that all the women’s “yes’s” are valued. While “communal” skills are more frequently mentioned in recommendation letters for promotion among women^31^, such efforts should receive more attention as research activities in evaluations. Further, women are more likely to experience imposter syndrome than men^73^, which may lead to a lack of confidence in publishing.

In SPH, there was a less clear trend of publication rates by ethnicity – there was an ethnicity effect only in women overall, and in men only when considering first/last author papers. However, with non-minoritized researchers having significantly more co-authors than minoritized researchers, this suggests that teamwork can be leveraged more by researchers from non-minoritized groups, potentially contributing to their productivity as measured in terms of publication count. This finding must be interpreted with caution as we did not have ethnicity recorded and the number of researchers of other ethnicities is low, such that an effect may not be detectable. Researchers of minority groups may have fewer opportunities for collaboration as very rarely afforded the opportunity to do so. We suggest further research on this.

The absence of a significant difference in overall publication rates between researchers from minoritized and non-minoritized groups implies that additional systemic barriers hinder the promotion of individuals from minoritized groups into tenure-track positions. In part, longer acceptance delay, fewer citation counts and underrepresentation on editorial boards are possibly contributing to the hurdles experienced by researchers from minoritized groups (and women). ^74,75^ Further, our findings suggest that there may be barriers to co-authoring papers led by first or last authors from minoritized groups such that they benefit less from the work effort of bigger teams, and, thus, produce fewer publications. The finding is important to discuss further as the overall proportion of researchers from minoritized groups is significantly smaller compared to London’s ethnicity profile, where ICL is based.^76^ Yet, the proportion of researchers from minoritized groups is similar to the proportion of people from minoritized groups living in England and Wales. ^77^ There may also be intersection with other factors affecting academic promotion. For example, in North America parental socioeconomic status has been shown to impact academic experiences^78^ and accessibility to tenure track positions, with tenure track holders nearly twice as likely to have PhD-holding parents as their non-tenure PhD-holding peers.^79^

To better understand why gender bias continues to exist in SPH, and to capture diverse insights and experiences which can inform a positive change in practise, we are running focus group discussions with representatives from all gender groups, ethnicity groups, and job levels. We also plan to expand this analysis beyond SPH to the broader Faculty of Medicine to investigate the presence of broader systemic bias and if there are groups within ICL that SPH can learn from.

We propose a more proactive approach to monitoring publication rates and other types of research engagement activities in faculties. We appreciate that the issue of faculty diversity in higher education has gained significant attention in recent years, especially in the US ^80^, and the literature on this matter is growing. But just as staff numbers are regularly analysed for the Athena Swan Charter, we advocate for routine, potentially automated, analyses of research engagement activities stratified by job category, ethnicity, and gender. This monitoring should be assigned as a paid responsibility to a designated individual or team, rather than relying on volunteers. Notably, the methodology should be chosen with respect to the field, appreciating that different fields have distinct research cultures. For example, natural sciences tend to prioritize collaborative journal articles while many humanities disciplines tend to value single-authored books or individual book chapters. This diversity challenges one-size-fits-all evaluation methods. Yet, our methodology and code may be used as a starting point for EDI in other settings.

Identifying and acknowledging biases is a crucial first step towards EDI. Departmental analysis, as used to inform this Commentary, can confirm existing hypotheses and guide focused action, while also revealing areas where investment may not be necessary. Failing to recognize bias has detrimental effects, but falsely assuming sexism is widespread in academic science also carries significant costs. It can discourage women from pursuing careers in scientific academia and lead to resources being misspent on addressing non-existent bias.^21^

Analyses and discussions as presented here are equally relevant for establishing more gender-balanced work environments in young, diverse institutions, and retaining diverse talent in faculties with a long history of inequities. For example, the Centre for Epidemiological Modelling and Analysis (CEMA), a recently founded (2020) Kenyan infectious disease modeling center has a female dominated infectious disease research environment (59% women). This example demonstrates that the pursuit of gender balance is an ongoing process, even in newly formed institutions. While some struggle with underrepresentation, others, like CEMA, may see an overrepresentation of women. Ultimately, achieving equity requires continuous effort and a commitment to inclusivity across all genders and ethnicities in all job levels.

It is our view that when addressing barriers to academia and promotion, we must ensure firstly that there is equity in opportunities to publish and secondly that research contributions other than publications and other work activities in which researchers may engage more are equally valued and recognized. Continued efforts should be made to achieve an inclusive research culture by examining potential biases in research funding, mentorship, and promotion processes, to support equity and retain a diverse generation of future researchers. We conclude this work with a set of recommendations for others to consider in evaluating EDI in their department (Text box 1).

## Ethics statement

This work did not require ethical approval from a human subject or animal welfare committee.

## Data accessibility statement

The code required to rerun the analysis in this study is available at https://github.com/paulachristen/publication_bias_script.

## Declaration of AI use

We have not used AI-assisted technologies in creating this article.

## Authors’ contributions statement

MB, TG, SL, MP, SR, NS: Data analysis; CM: Data extraction; AA, SH, WK, JH: conception of further research; KM, PC, JM, TN, HT, AC, IB, LCO, CU, SvE, DF, GC, SEQ: Study conception, analysis design, writing.

## Funding statement

The authors acknowledge funding for this project from Imperial College London’s School of Public Health, the National Institute for Health and Care Research (NIHR) Health Protection Research Unit in Modelling and Health Economics, a partnership between the UK Health Security Agency, Imperial College London and LSHTM (grant code NIHR200908) and the MRC Centre for Global Infectious Disease Analysis (reference MR/X020258/1), funded by the UK Medical Research Council (MRC). This UK funded award is carried out in the frame of the Global Health EDCTP3 Joint Undertaking. Disclaimer: *“The views expressed are those of the author(s) and not necessarily those of the NIHR, UK Health Security Agency or the Department of Health and Social Care.”* LCO acknowledges funding from the UK Royal Society. SEQ acknowledges funding from the Wellcome Institutional Strategic Support Fund (ISSF).

### [Text box] Recommendations

1. **Regular Monitoring and Analysis:** Implement routine, automated, analyses of research engagement activities to account for nuances in productivity measures, stratified by job category, ethnicity, and gender using empirical data (i.e., ideally not relying on probabilistically assigned ethnicity information). This monitoring should be an assigned, paid responsibility (e.g., individuals at EDI Centres), not reliant on volunteers, and should be reviewed regularly at departmental meetings (e.g., Athena Swan and Race Equality Charter committee meetings, senior management meetings, people and culture committees).
2. **Mixed-Methods Approach:** Utilize both qualitative and quantitative data for comprehensive evaluations of barriers and facilitators to publishing, career progression, and maintaining a diverse and accepting working environment, drawing on the strengths of each approach.
3. **What Works:** Determining what institutional environments and policies reduce publication bias to draw up a comprehensive, clear list of recommendations that departments can act on.
4. **Early Intervention in Young Institutions:** Proactively establish equitable work environments and policies in newly founded institutions to prevent the entrenchment of biases.
5. **Acknowledge and Address Biases:** Openly identify and acknowledge biases as well as successes within the system and commit to focused action to address and maintain them.

## Data Availability

All data prduced in the present study are available upon reasonable request to the authors.

https://github.com/paulachristen/publication_bias_script

## Supplement

### A. Methods

#### Data collection

##### Publications

Data on publications were downloaded through the research information management system, Symplectic Elements. The system automatically identifies new publications as they become available online and alerts staff members to claim authorship which links output to their personal profiles. Individuals can also manually add research activities into the database. Data retrieved via Symplectic Elements included the name of the staff member linked to each publication, co-author names, publication title, publication date and publication type.

##### Gender

Each SPH author was assigned an assumed gender using the gender (v0.6.0^48^) and genderdata (v0.6.0^81^) R packages which inferred gender based on probabilities of a name being men or women in historical datasets. For this research we used a method using names from the U.S. Social Security Administration for babies born between 1940 and 2000. A match was made when the probability of a name being associated with either a men or women was greater than 80%. When the probability was lower than 80%, the gender was determined from a web search.

##### Job category

Each individual was also assigned a job category by scraping data from ICL staff webpages using the rvest (v1.0.3^82^) package. As information on promotion dates were not readily available, we relied on the assumption that the most recent SPH job title was an individual’s job category across all publication records. Individuals with a missing job category were reviewed manually. Anyone who was not ascribed the job title of Student / Research Assistant, Research Associate, Research Fellow, Lecturer, Senior Lecturer / Reader or Professor was classified as ‘Other’. ‘Others’ refers to individuals, who assume or hold administrative/managerial roles, honorary affiliations, visiting researcher status, temporary (casual worker) positions, clinical roles or trial/funding specific job titles.

##### Ethnicity

Ethnicity was assigned using the NamePrism^49^ Application Programming Interface which generated probabilistic estimates of individual ethnicity based on SPH author last name using 2020 data from the U.S. Census Bureau. Ethnicities were categorized based on the predicted probability of the last name being white, black, Hispanic, Asian, or other, with ethnicity assigned to the category with the maximum probability.

#### Data

30,252 publication records linked to 690 staff at SPH were downloaded from Symplectic Elements, including all claimed publications from the period January 1, 2014, to June 5, 2023.

108 (15.7%) individuals were not matched to gender, 94 due to a missing assignment and 14 due to a low probability of a correct match. Individuals with a missing gender assignment were reviewed and assigned manually. Individuals with a missing job category (n = 87) were reviewed manually.

Publication entries were removed if they fell under the following categories: abstracts, reprints, preprints, book reviews, news items, retractions, corrections, erratum, and PhD theses (n = 6,557 / 30,252; 21.7%). Entries were removed if they contained work published prior to 2014 (n = 19) or were missing a publication date (n = 93). Entries from honorary or visiting staff positions were also removed (n = 4,244). Following data cleaning, 18,322 records remained.

From 1 January 2014 to 5 June 2023, 18,322 publication records were authored and claimed by 528 SPH staff and students, 279 (52.8%) of whom were classified as woman and 249 (47.2%) as men. Of all publications, 5,776 were claimed by women (29.9%) and 13,563 by men (70.1%). Women had a median of 6 publications (IQR: 2-20) and men had a median of 15 publications (IQR: 5-62). Women authored publications with a median of 9 (IQR: 6-18) authors and men with a median of 10 (IQR: 6-23) authors.

Of all first-author publications (n = 1,919), 785 (40.9%) were linked to women and 1,134 (59.1%) were linked to men. Of all last-author publications (n = 3,362), 804 (23.9%) were linked to women and 2,558 (76.1%) were linked to men.

#### Data analysis

We fitted negative binomial mixed-effects regression models to assess the effects of time, gender, ethnicity, and job level on total, first or last author, and middle author publications per year using the glmmTMB (v1.1.9^83^) package. Based on internal co-authorships, we performed a network analysis to quantify every author’s centrality to the department’s publication network and estimated the association between gender, ethnicity, and the authors’ number of internal co-authors.

We estimated the publishing years gap between women and men using unadjusted incidence rate ratios (IRRs). The publication rate stratified by gender and job seniority was calculated by dividing the total number of publications by the total number of person-years. To determine person-years, we assumed that researchers were active every year between their first and last publication. The publication gap was calculated using: (1 - 1/IRR) * 10, where 10 represents the number of years in the study period (2014–2023).

### B. Support

**Figure 2.**
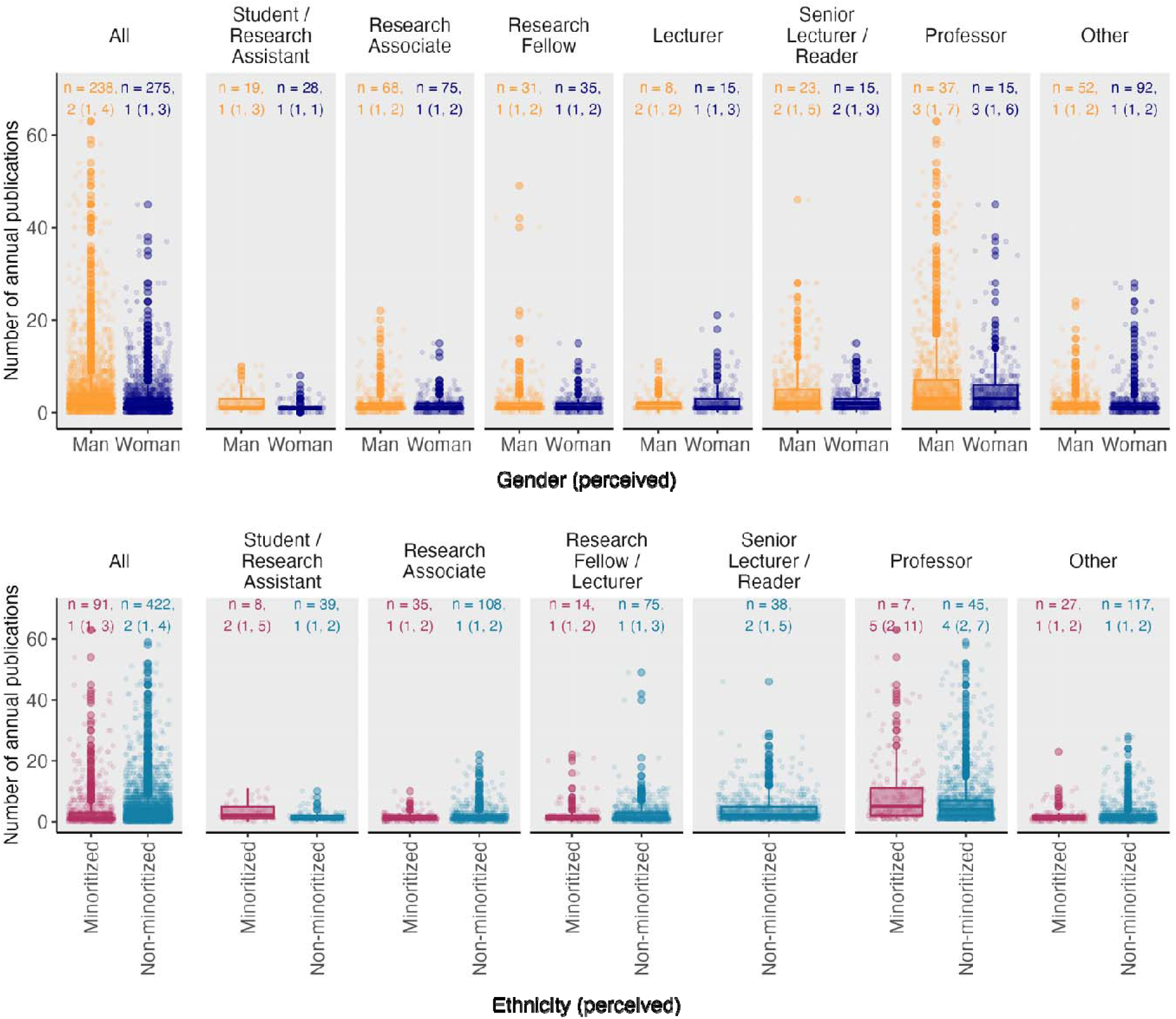
Mean number of annual publications by job category, ethnicity (perceived) and gender (perceived). The box represents the median and interquartile range (IQR) of mean number of annual publications. The boxplot whiskers represent the adjacent values, which are by convention within 1.5 times the interquartile range. The dots represent mean number of annual publications outside of the adjacent values, known as outliers. n = number of observations. Annotation on graph: median (IQR). See Data Collection (job category) section for more information on “other”.

**Figure 3.**
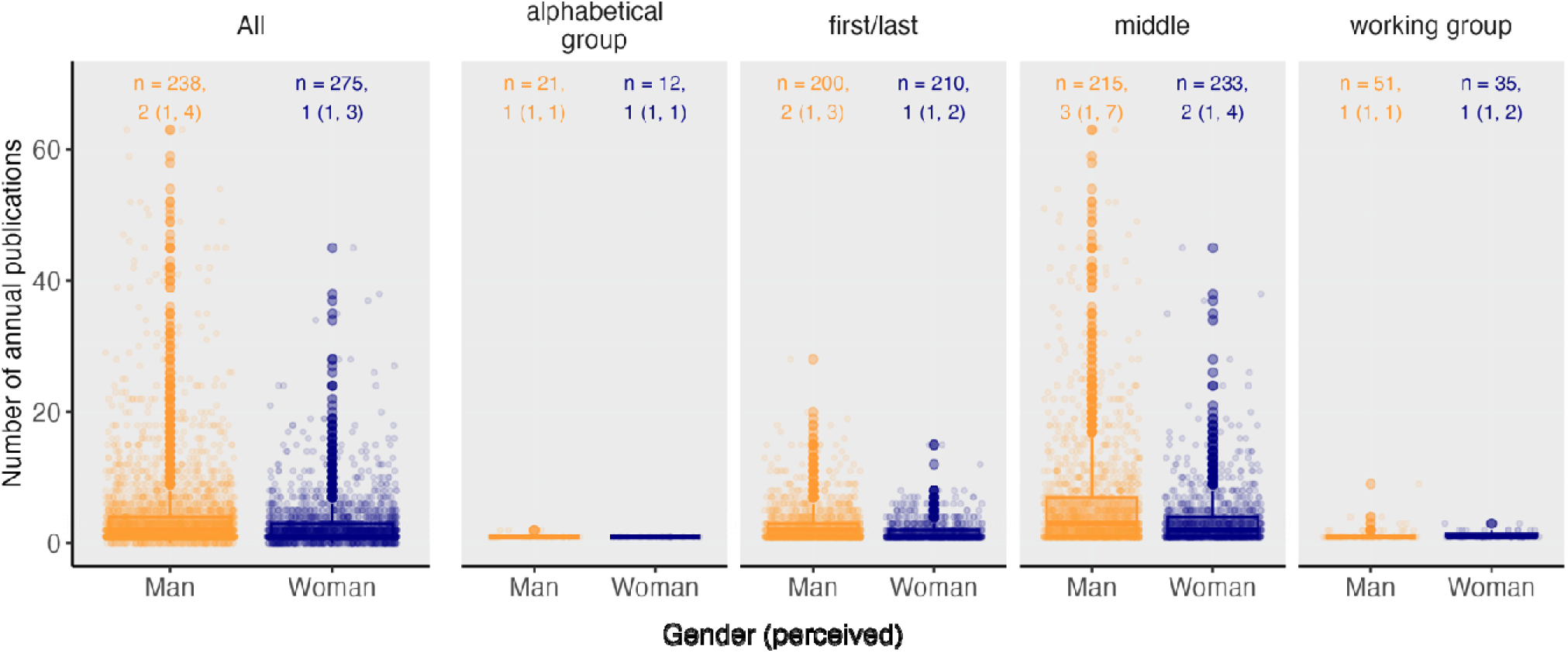
Mean number of annual publications by authorship position, ethnicity (perceived) and gender (perceived). The box represents the median and interquartile range (IQR) of mean number of annual publications. The boxplot whiskers represent the adjacent values, which are by convention within 1.5 times the interquartile range. The dots represent mean number of annual publications outside of the adjacent values, known as outliers. n = number of observations. Annotation on graph: median (IQR). See Data Collection (job category) section for more information on “other”.

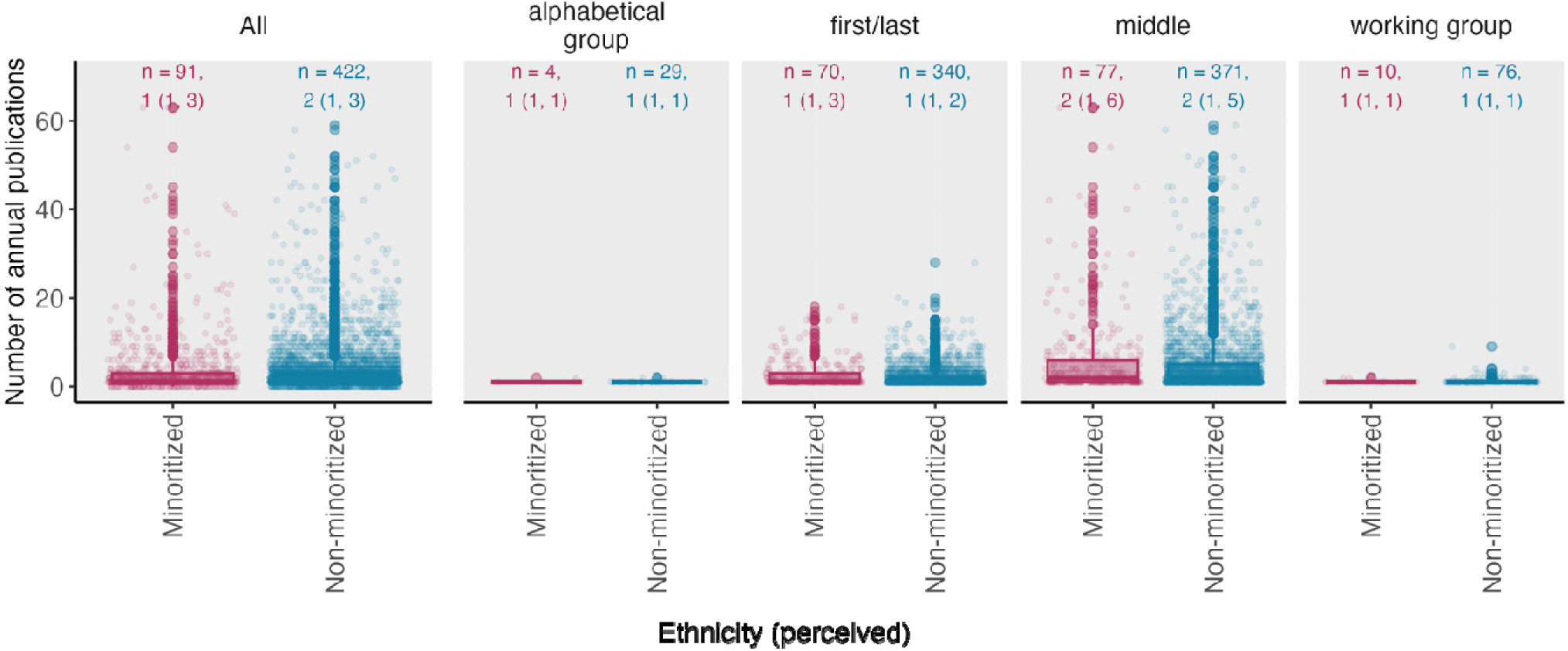

**Table 1.** Negative binomial mixed-effects regression analysis assessing the predictive factors of mean annual publication count. IRR: incidence rate ratio; 95% CI: 95% confidence interval. *p < 0.05, **p < 0.01, ***p < 0.001.

|  | All annual publications per person |  | First/last authored annual publications per person |  | Middle authored annual publications per person |  | Internal co-authors per year |  |
| --- | --- | --- | --- | --- | --- | --- | --- | --- |
| Predictors | IRR | CI | IRR | CI | IRR | CI | IRR | CI |
| Intercept | 1.74 *** | 1.31 – 2.31 | 0.63 * | 0.42 – 0.94 | 0.89*** | 0.62 - 1.28 | 0.60 * | 0.38 – 0.96 |
| Gender (n = 513) |  |  |  |  |  |  |  |  |
| Woman (n = 275) | 1 |  | 1 |  | 1 |  | 1 |  |
| Man (n = 238) | 1.30 *** | 1.15 – 1.46 | 1.31 *** | 1.12 – 1.53 | 1.35*** | 1.16 - 1.57 | 1.01 | 0.82 – 1.25 |
| Ethnicity (perceived) (n = 513) |  |  |  |  |  |  |  |  |
| Minoritized (n = 91) | 1 |  | 1 |  | 1 |  | 1 |  |
| Non-minoritized (n = 422) | 1.05 | 0.89 – 1.23 | 0.95 | 0.77 – 1.18 | 1.07 | 0.87 - 1.32 | 1.81 *** | 1.35 – 2.42 |
| Time |  |  |  |  |  |  |  |  |
| 2014 – 2015 (n = 246) | 1 |  | 1 |  | 1 |  | 1 |  |
| 2016 – 2017 (n = 272) | 1.04 | 0.95 – 1.14 | 0.99 | 0.87 – 1.12 | 1.10 | 0.99 - 1.24 | 1.25 *** | 1.10 – 1.43 |
| 2018 – 2019 (n = 331) | 1.11 * | 1.02 – 1.22 | 1.05 | 0.93 – 1.18 | 1.24 *** | 1.11 - 1.38 | 1.62 *** | 1.43 – 1.85 |
| 2020 – 2021 (n = 410) | 1.18 *** | 1.08 – 1.28 | 0.93 | 0.83 – 1.05 | 1.37 *** | 1.24 - 1.52 | 3.79 *** | 3.37 – 4.27 |
| 2022 – 2023 (n = 377) | 0.73 *** | 0.66 – 0.79 | 0.60 *** | 0.53 – 0.68 | 0.83 ** | 0.75 - 0.93 | 2.11 *** | 1.86 – 2.39 |
| Job Category (n = 513) |  |  |  |  |  |  |  |  |
| Student / Research Assistant (n = 47) | 1 |  | 1 |  | 1 |  | 1 |  |
| Research Associate (n = 143) | 0.96 | 0.74 – 1.24 | 0.99 | 0.68 – 1.43 | 0.92 | 0.66 - 1.29 | 0.93 | 0.61 – 1.42 |
| Research Fellow (n = 66) | 1.21 | 0.92 – 1.61 | 1.35 | 0.92 – 1.99 | 1.21 | 0.85 - 1.73 | 1.16 | 0.73 – 1.85 |
| Lecturer (n = 23) | 1.67 ** | 1.19 – 2.34 | 1.83 ** | 1.16 – 2.88 | 1.70 * | 1.1 - 2.61 | 0.87 | 0.48 – 1.58 |
| Senior Lecturer/Reader (n = 38) | 2.79 *** | 2.08 – 3.76 | 2.72 *** | 1.82 – 4.07 | 3.06 *** | 2.1 - 4.45 | 2.42 *** | 1.45 – 4.04 |
| Professor (n = 52) | 5.43 *** | 4.10 – 7.18 | 5.10 *** | 3.48 – 7.48 | 5.75 *** | 4.03 - 8.19 | 4.22 *** | 2.62 – 6.81 |
| Other (n = 144) | 1.09 | 0.84 – 1.41 | 0.89 | 0.61 – 1.28 | 1.08 | 0.78 - 1.50 | 0.80 | 0.53 – 1.23 |
| Random Effects |  |  |  |  |  |  |  |  |
| $\sigma^2$ | 0.50 | | 0.00 | | 0.00 | | 0.81 | |
| $\tau_{00}$ | 0.27<sub>Username</sub> | | 0.40<sub>Username</sub> | | 0.44<sub>Username</sub> | | 1.08<sub>Username</sub> | |
| ICC | 0.35 |  | 1.00 |  | 1.00 |  | 1.00 |  |
| N | 513<sub>Username</sub> |  | 513<sub>Username</sub> |  | 513<sub>Username</sub> |  | 513<sub>Username</sub> |  |
| Observations | 3025 |  | 3025 |  | 3025 |  | 3025 |  |
| Marginal R<sup>2</sup> / Conditional R<sup>2</sup> | 0.407 / 0.614 |  | 0.581 / 1.000 |  | 0.571 / 1.000 |  | 0.374 / 1.000 |  |
| AIC | 14747.448 |  | 9268.875 |  | 12959.474 |  | 15147.519 |  |

**Table 2.**
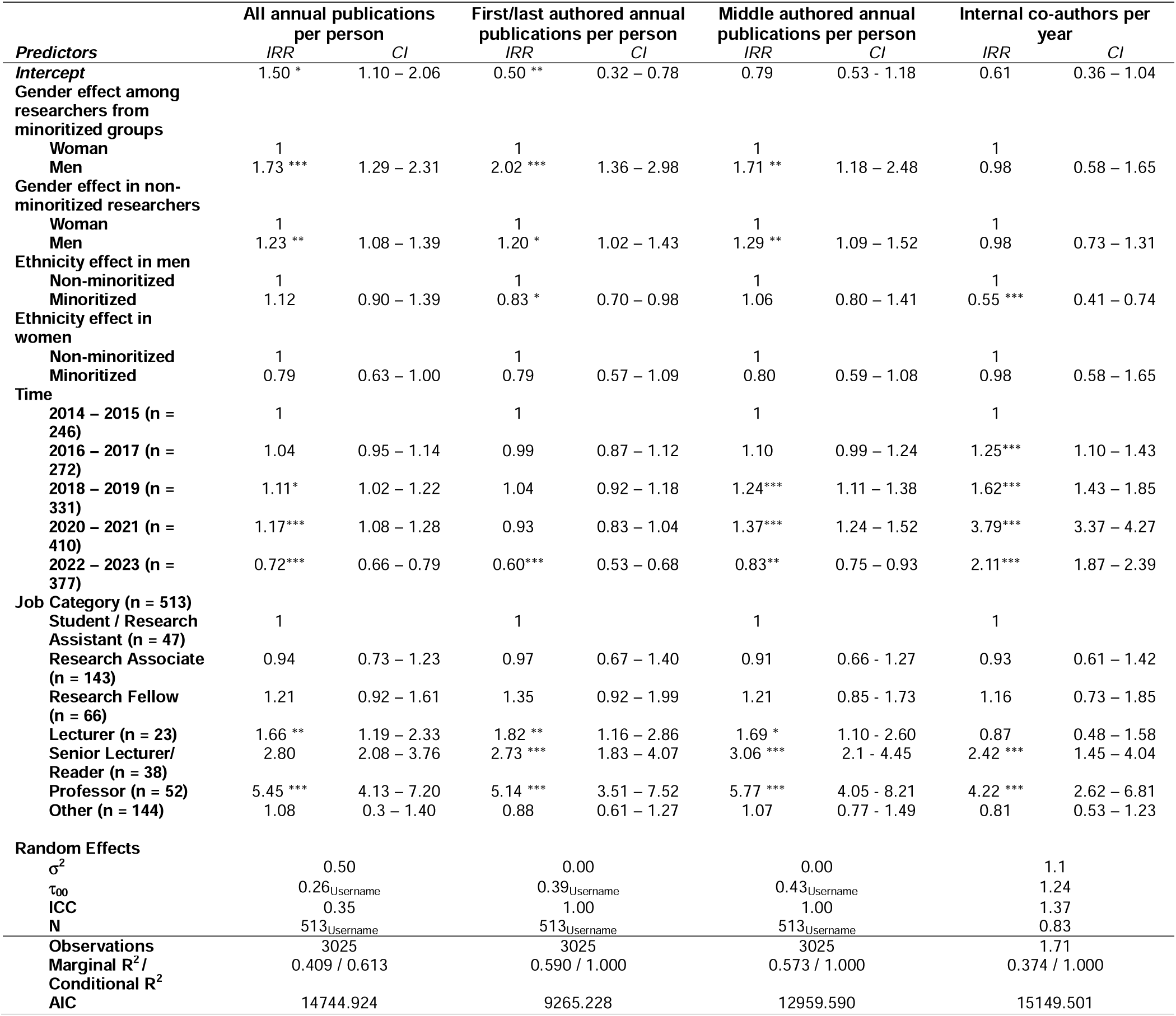
Negative binomial mixed-effects regression analysis assessing the predictive factors of mean annual publication count, including interaction terms between ethnicity and gender in the model. IRR: incidence rate ratio; 95% CI: 95% confidence interval. *p < 0.05, **p < 0.01, ***p < 0.001.

| Predictors | All annual publications per person |  | First/last authored annual publications per person |  | Middle authored annual publications per person |  | Internal co-authors per year |  |
| --- | --- | --- | --- | --- | --- | --- | --- | --- |
|  | IRR | CI | IRR | CI | IRR | CI | IRR | CI |
| Intercept | 1.50 * | 1.10 – 2.06 | 0.50 ** | 0.32 – 0.78 | 0.79 | 0.53 – 1.18 | 0.61 | 0.36 – 1.04 |
| Gender effect among researchers from minoritized groups |  |  |  |  |  |  |  |  |
| Woman | 1 |  | 1 |  | 1 |  | 1 |  |
| Men | 1.73 *** | 1.29 – 2.31 | 2.02 *** | 1.36 – 2.98 | 1.71 ** | 1.18 – 2.48 | 0.98 | 0.58 – 1.65 |
| Gender effect in non-minoritized researchers |  |  |  |  |  |  |  |  |
| Woman | 1 |  | 1 |  | 1 |  | 1 |  |
| Men | 1.23 ** | 1.08 – 1.39 | 1.20 * | 1.02 – 1.43 | 1.29 ** | 1.09 – 1.52 | 0.98 | 0.73 – 1.31 |
| Ethnicity effect in men |  |  |  |  |  |  |  |  |
| Non-minoritized | 1 |  | 1 |  | 1 |  | 1 |  |
| Minoritized | 1.12 | 0.90 – 1.39 | 0.83 * | 0.70 – 0.98 | 1.06 | 0.80 – 1.41 | 0.55 *** | 0.41 – 0.74 |
| Ethnicity effect in women |  |  |  |  |  |  |  |  |
| Non-minoritized | 1 |  | 1 |  | 1 |  | 1 |  |
| Minoritized | 0.79 | 0.63 – 1.00 | 0.79 | 0.57 – 1.09 | 0.80 | 0.59 – 1.08 | 0.98 | 0.58 – 1.65 |
| Time |  |  |  |  |  |  |  |  |
| 2014 – 2015 (n = 246) | 1 |  | 1 |  | 1 |  | 1 |  |
| 2016 – 2017 (n = 272) | 1.04 | 0.95 – 1.14 | 0.99 | 0.87 – 1.12 | 1.10 | 0.99 – 1.24 | 1.25*** | 1.10 – 1.43 |
| 2018 – 2019 (n = 331) | 1.11* | 1.02 – 1.22 | 1.04 | 0.92 – 1.18 | 1.24*** | 1.11 – 1.38 | 1.62*** | 1.43 – 1.85 |
| 2020 – 2021 (n = 410) | 1.17*** | 1.08 – 1.28 | 0.93 | 0.83 – 1.04 | 1.37*** | 1.24 – 1.52 | 3.79*** | 3.37 – 4.27 |
| 2022 – 2023 (n = 377) | 0.72*** | 0.66 – 0.79 | 0.60*** | 0.53 – 0.68 | 0.83** | 0.75 – 0.93 | 2.11*** | 1.87 – 2.39 |
| Job Category (n = 513) |  |  |  |  |  |  |  |  |
| Student / Research Assistant (n = 47) | 1 |  | 1 |  | 1 |  | 1 |  |
| Research Associate (n = 143) | 0.94 | 0.73 – 1.23 | 0.97 | 0.67 – 1.40 | 0.91 | 0.66 – 1.27 | 0.93 | 0.61 – 1.42 |
| Research Fellow (n = 66) | 1.21 | 0.92 – 1.61 | 1.35 | 0.92 – 1.99 | 1.21 | 0.85 – 1.73 | 1.16 | 0.73 – 1.85 |
| Lecturer (n = 23) | 1.66 ** | 1.19 – 2.33 | 1.82 ** | 1.16 – 2.86 | 1.69 * | 1.10 – 2.60 | 0.87 | 0.48 – 1.58 |
| Senior Lecturer/Reader (n = 38) | 2.80 | 2.08 – 3.76 | 2.73 *** | 1.83 – 4.07 | 3.06 *** | 2.1 – 4.45 | 2.42 *** | 1.45 – 4.04 |
| Professor (n = 52) | 5.45 *** | 4.13 – 7.20 | 5.14 *** | 3.51 – 7.52 | 5.77 *** | 4.05 – 8.21 | 4.22 *** | 2.62 – 6.81 |
| Other (n = 144) | 1.08 | 0.3 – 1.40 | 0.88 | 0.61 – 1.27 | 1.07 | 0.77 – 1.49 | 0.81 | 0.53 – 1.23 |
| Random Effects |  |  |  |  |  |  |  |  |
| $\sigma^2$ | 0.50 | | 0.00 | | 0.00 | | 1.1 | |
| $\tau_{00}$ | 0.26<sub>Username</sub> | | 0.39<sub>Username</sub> | | 0.43<sub>Username</sub> | | 1.24 | |
| ICC | 0.35 |  | 1.00 |  | 1.00 |  | 1.37 |  |
| N | 513<sub>Username</sub> |  | 513<sub>Username</sub> |  | 513<sub>Username</sub> |  | 0.83 |  |
| Observations | 3025 |  | 3025 |  | 3025 |  | 1.71 |  |
| Marginal R<sup>2</sup> / Conditional R<sup>2</sup> | 0.409 / 0.613 |  | 0.590 / 1.000 |  | 0.573 / 1.000 |  | 0.374 / 1.000 |  |
| AIC | 14744.924 |  | 9265.228 |  | 12959.590 |  | 15149.501 |  |

**Table 3.** Negative binomial mixed-effects regression analysis assessing the predictive factors of mean annual publication count, including interaction terms between gender and time period in the model. IRR: incidence rate ratio; 95% CI: 95% confidence interval. *p < 0.05, **p < 0.01, ***p < 0.001.

| Predictors | All annual publications per person |  | First/last authored annual publications per person |  | Middle authored annual publications per person |  | Internal co-authors per year |  |
| --- | --- | --- | --- | --- | --- | --- | --- | --- |
|  | IRR | CI | IRR | CI | IRR | CI | IRR | CI |
| Intercept | 1.76 *** | 1.31 – 2.36 | 0.65 * | 0.43 – 0.98 | 0.88 | 0.61 – 1.29 | 0.62 * | 0.38 – 1.00 |
| Time effect among men (n = 275) |  |  |  |  |  |  |  |  |
| 2014 – 2015 (n = 132) | 1 |  | 1 |  | 1 |  | 1 |  |
| 2016 – 2017 (n = 145) | 1.13 | 0.93 – 1.36 | 1.03 | 0.79 – 1.34 | 1.21 | 0.95 – 1.53 | 1.13 | 0.86 – 1.49 |
| 2018 – 2019 (n = 168) | 1.22 * | 1.02 – 1.47 | 1.36 * | 1.06 – 1.76 | 1.15 | 0.92 – 1.45 | 1.21 | 0.93 – 1.57 |
| 2020 – 2021 (n = 202) | 0.98 | 0.83 – 1.17 | 1.03 | 0.81 – 1.30 | 0.93 | 0.75 – 1.15 | 1.06 | 0.83 – 1.35 |
| 2022 – 2023 (n = 195) | 0.83 * | 0.69 – 1.00 | 0.78 | 0.61 – 1.01 | 0.81 | 0.65 – 1.02 | 0.88 | 0.69 – 1.14 |
| Time effect among women (n = 238) |  |  |  |  |  |  |  |  |
| 2014 – 2015 (n = 114) | 1 |  | 1 |  | 1 |  | 1 |  |
| 2016 – 2017 (n = 127) | 0.89 | 0.73 – 1.07 | 0.97 | 0.75 – 1.26 | 0.83 | 0.65 – 1.05 | 0.88 | 0.67 – 1.16 |
| 2018 – 2019 (n = 163) | 0.82 * | 0.68 – 0.98 | 0.73 * | 0.57 – 0.95 | 0.87 | 0.69 – 1.09 | 0.83 | 0.64 – 1.08 |
| 2020 – 2021 (n = 208) | 1.02 | 0.86 – 1.21 | 0.97 | 0.77 – 1.23 | 1.08 | 0.87 – 1.33 | 0.95 | 0.74 – 1.20 |
| 2022 – 2023 (n = 182) | 1.21 * | 1.00 – 1.45 | 1.28 | 0.99 – 1.65 | 1.23 | 0.98 – 1.55 | 1.13 | 0.88 – 1.46 |
| Gender (n = 513) |  |  |  |  |  |  |  |  |
| Woman (n = 275) | 1 |  | 1 |  | 1 |  | 1 |  |
| Man (n = 238) | 1.29 ** | 1.08 – 1.54 | 1.28 * | 1.01 – 1.62 | 1.38 ** | 1.1 – 1.73 | 0.98 | 0.73 – 1.31 |
| Ethnicity (perceived) (n = 513) |  |  |  |  |  |  |  |  |
| Minoritized (n = 91) | 1 |  | 1 |  | 1 |  | 1 |  |
| Non-minoritized (n = 422) | 1.04 | 0.89 – 1.23 | 0.95 | 0.77 – 1.18 | 1.07 | 0.87 – 1.31 | 1.81 *** | 1.35 – 2.42 |
| Job Category (n = 513) |  |  |  |  |  |  |  |  |
| Student / Research Assistant (n = 47) | 1 |  | 1 |  | 1 |  |  |  |
| Research Associate (n = 143) | 0.95 | 0.73 – 1.24 | 0.97 | 0.67 – 1.40 | 0.92 | 0.66 – 1.29 | 0.92 | 0.60 – 1.41 |
| Research Fellow (n = 66) | 1.21 | 0.92 – 1.60 | 1.34 | 0.91 – 1.97 | 1.22 | 0.85 – 1.73 | 1.16 | 0.72 – 1.85 |
| Lecturer (n = 23) | 1.66 ** | 1.18 – 2.33 | 1.80 * | 1.15 – 2.84 | 1.70 * | 1.11 – 2.62 | 0.86 | 0.47 – 1.58 |
| Senior Lecturer/Reader (n = 38) | 2.78 *** | 2.07 – 3.74 | 2.69 *** | 1.80 – 4.02 | 3.06 *** | 2.1 – 4.46 | 2.41 *** | 1.44 – 4.03 |
| Professor (n = 52) | 5.40 *** | 4.08 – 7.14 | 5.04 *** | 3.44 – 7.38 | 5.75 *** | 4.03 – 8.19 | 4.19 *** | 2.60 – 6.77 |
| Other (n = 144) | 1.09 | 0.84 – 1.41 | 0.88 | 0.61 – 1.27 | 1.08 | 0.78 – 1.51 | 0.80 | 0.52 – 1.23 |
| Random Effects |  |  |  |  |  |  |  |  |
| $\sigma^2$ | 0.50 | | 0.00 | | 0.00 | | 0.00 | |
| $\tau_{00}$ | 0.27<sub>Username</sub> | | 0.39<sub>Username</sub> | | 0.44<sub>Username</sub> | | 1.09<sub>Username</sub> | |
| ICC | 0.35 |  | 1.00 |  | 1.00 |  | 1.00 |  |
| N | 513<sub>Username</sub> |  | 513<sub>Username</sub> |  | 513<sub>Username</sub> |  | 513<sub>Username</sub> |  |
| Observations | 3025 |  | 3025 |  | 3025 |  | 3025 |  |
| Marginal R<sup>2</sup> / Conditional R<sup>2</sup> | 0.408 / 0.615 |  | 0.582 / 1.000 |  | 0.572 / 1.000 |  | 0.374 / 1.000 |  |
| AIC | 14732.068 |  | 9256.280 |  | 12949.173 |  | 1.5146.189 |  |

